# The Effect of the COVID-19 Pandemic on Life Expectancy in the US: An Application of Hybrid Life Expectancy

**DOI:** 10.1101/2022.12.19.22283697

**Authors:** Warren C. Sanderson, Sergei Scherbov

## Abstract

Life expectancy at birth (both sexes) in the US was 1.9 years lower in 2020 than in 2019. This reduction has been widely discussed. This 1.9 year reduction compares a group of people who would live their entire lives at the 2019 survival rates with a group that would live their entire lives experiencing the pandemic survival rates of 2020. Here we introduce hybrid life expectancy. This is the life expectancy of people who experience the 2020 survival rates for various shorter periods followed by a return to the 2019 levels. For pandemic durations of 10 years or less life expectancy at birth does not fall from its 2019 level. For a pandemic duration of 10 years, the life expectancy of 65 year-olds would fall by 0.2 years.

## Introduction

According to the Human Mortality Database (HMD), life expectancy at birth (both sexes) in the United States fell from 78.99 years in 2019 to 77.07 years in 2020. (1) A decrease of 1.9 years in life expectancy is certainly noteworthy and it has been discussed widely. (2–4) The 1.9 year reduction of in life expectancy at birth compares the life expectancy of two hypothetical groups of people, one of whom live their lives experiencing the baseline survival rates of 2019 and the other of whom live their entire lives experiencing the lower SARS-CoV-2-affected survival rates of 2020. Life expectancy calculated in this way is called period life expectancy because it assumes that people life their entire lives experiencing the survival rates of a specific period. In particular, the 2020 period life expectancy at birth assumes that people life their entire lives in a never ending pandemic.

In order to assess the effects of a pandemic of limited duration on life expectancy, we introduce hybrid life expectancy. The hybrid life expectancy of people assumes that they experience pandemic-level survival rates only for a fixed number of years after which survival rates return to their baseline level. In this paper, we contrast the difference in life expectancy reductions, assuming that people live their entire lives with pandemic survival rates and assuming that the pandemic is limited in its duration.

## Methods

The survival rates that we use come from the Human Mortality Database. (1) These data differ slightly from those published by the Centers for Disease Control and Prevention (CDC). For example, the CDC reports that the decrease between 2019 and 2020 in period life expectancy at birth (both sexes) was years, (5) 0.1 years different from the HMD figure. Small differences like these can result from a variety of methodological details. We use HMD data here because, at this writing, single year of age survival rates are available there.

Table 1 shows 1-year survival rates for selected ages during 2019 and the COVID year 2020.

**Table 1:** 1-year survival rates (both sexes) for selected ages, 2019 and 2020.

| Age | Year |  |
| --- | --- | --- |
|  | 2019 | 2020 |
| 5 | 0.99987 | 0.99987 |
| 45 | 0.99737 | 0.99677 |
| 65 | 0.98732 | 0.98512 |
| 80 | 0.95430 | 0.94582 |

Source: Human Morality Database (1)

In 2019, the 1-year survival rate of 5-year-olds was 0.99987. During 2020 the rate was also 0.99987. The pandemic had no effect on the survival rates of 5-year-olds. For 80-year-olds the survival rate fell from 0.95430 to 0.94582.

Hybrid period life expectancy is a hypothetical construct that integrates the survival rates of two periods, a pandemic period and a baseline period that follows. For example, the hybrid life expectancy of a 50-year-old who experienced a 5 year long COVID pandemic would be computed first using the 2020 survival rates for people 50 through 54 and then 2019 survival rates for the rest of their lives.

## Results

Our results are shown in Table 2. The table shows the difference between period life expectancy in 2019 and the hybrid life expectancy for selected ages and pandemic durations. We assume that in each pandemic year survival rates are the same as in 2020. When the duration of the pandemic is the entire lifetime, the table shows the difference between period life expectancy in 2020 and period life expectancy in 2019.

**Table 2:** Life Expectancy in 2019 and Reductions due to the Pandemics of Various Duration, Selected Ages (both sexes)

| PANDEMIC<br>DURATION | Age |  |  |  |
| --- | --- | --- | --- | --- |
|  | At Birth | 20 | 65 | 80 |
| 5 | 0 | 0 | 0.2 | 0.4 |
| 10 | 0 | 0.1 | 0.4 | 0.7 |
| Whole life | 1.9 | 1.9 | 1.2 | 0.9 |
| Baseline (2019) life expectancy | 79.0 | 59.7 | 19.7 | 9.5 |
| Pandemic (2020) life expectancy | 77.1 | 57.8 | 18.5 | 8.6 |
| Hybrid life expectancy (duration 1 year) | 79.0 | 59.7 | 19.7 | 9.4 |

Sources: Human Mortality Database (1) and authors' computations
Note: Life expectancy reductions are computed as life expectancy in 2019 minus the corresponding hybrid life expectancy.

Table 2 shows that for pandemic durations of 10 years or less there is no reduction in life expectancy at birth. A ten year long pandemic would affect children in the first ten years of their lives, and their survival rates in 2020 were almost identical to those in 2019. Even 20-year-olds, experiencing a decade of reduced survival rates due to the pandemic, would lose only one-tenth of a year of life expectancy. Eighty-year-olds experience greater life expectancy reductions. In 2019, life expectancy at 80 (both sexes) was 9.5 years. If they experienced the 2020 survival rates from their 80th birthday to their 90th, their life expectancy would be reduced by 0.7 years. If pandemic survival rates were used for the rest of their lives instead of 10 years, the life expectancy reduction would be 0.9 years.

For pandemic durations of one year, life expectancy reductions would be less than one-tenth of a year for ages 65 and below. The overall patterns are similar for males and females.

## Discussion

The main advantage of our approach is that it takes the duration of the pandemic into account. Pandemics, by their nature, are time limited events. For example, if the reduced survival rates of 2020 were to last for five years, from 2020 though 2024, and survival rates were then to return permanently to their 2019 levels, the pandemic would have no effect on the life expectancy of people born in 2020. In the same scenario, the reduction in the life expectancy of 65-year-olds at the beginning of the pandemic would be 0.2 years. A measurement of the life expectancy difference for 65-year-olds, ignoring the pandemic’s duration, produces a figure of 1.2 years. When pandemics take place over multiple years life expectancy differences from a base year to any year in the pandemic do not reflect the experience of people.

Hybrid life expectancy can be computed after the pandemic has ended and its associated survival rates known. In this paper, we computed hybrid life expectancy while we are in the midst of the pandemic. We did this by assuming one pattern of reduced survival rates that persists for the duration of the pandemic. This could cause of problems if future pandemic survival rates differed widely from the assumed pattern. At this writing there is only one year of single-year-of-age pandemic survival rates available, those for 2020. Fortunately, the 2020 survival rates are a plausible choice. Life expectancy at birth was lower in 2020 than in 2019 and it was lower still in 2021. (5) Although the final data are not available at this time, it is likely, based on the number of COVID deaths, that life expectancy at birth in 2022 will be higher than the 2020 figure. (6) So 2020, not having the highest or lowest life expectancy at birth, is a plausible choice. In more complex cases, the choice of a single pattern of survival rates could be difficult.

It is not known when the current pandemic will end. If the pandemic lasts less than a decade, then the life expectancy reductions shown in Table 2 for a 10-year pandemic are likely to be upper bounds on the amounts that would be observed.

Pandemics are not all the same (7). They have different patterns of age-specific survival rates. The results in Table 2 are specific to the COVID-19 pandemic in the US. The results for pandemics at different times and places would be different, but the use of hybrid life expectancy would still be appropriate.

The reduction in life expectancy at birth between 2019 and 2020 of 1.9 years is not a mistake, but it can be easily misinterpreted. It does not mean that children born in 2020 would have an average of fewer years of life compared to those born in 2019 or that the average person would have lost an average of 1.9 years of life because of the reduced survival rates of 2020. Hybrid life expectancy reductions can also be misinterpreted. The observation that a pandemic of a decade or less had no effect on life expectancy at birth does not mean that the pandemic had no effect whatsoever. Pandemics with large numbers of deaths may also feature small life expectancy reductions. (8)

The life expectancy data and calculations used here assume that all members of the population have identical frailty. Hybrid life expectancy is a concept based on observed survival rates. A more complex approach to hybrid life expectancy could take differential frailty into account. (9,10) If the more frail die in a pandemic, the population remaining at the end of the pandemic would be more robust than those in the baseline population and, therefore, have higher survival rates. On the other hand, if infection with the SARS-CoV-2 virus caused lingering effects that reduced subsequent survival rates, those rates would be lower than in the baseline. Incorporating these effects could affect the results reported here.

## Data Availability

All data produced in the present work are contained in the manuscript.

https://www.mortality.org

## References

1. Max Planck Institute for Demographic Research (Germany), University of California, Berkeley (USA), French Institute for Demographic Studies (France). HMD. Human Mortality Database [Internet]. [cited 2022 Apr 4]. Available from: https://www.mortality.org

2. Andrasfay T, Goldman N. Reductions in 2020 US life expectancy due to COVID-19 and the disproportionate impact on the Black and Latino populations. Proceedings of the National Academy of Sciences [Internet]. 2021 Feb 2 [cited 2022 Aug 22];118(5):e2014746118. Available from: https://www.pnas.org/doi/10.1073/pnas.2014746118

3. Woolf SH, Masters RK, Aron LY. Changes in Life Expectancy Between 2019 and 2020 in the US and 21 Peer Countries. JAMA Network Open [Internet]. 2022 Apr 13 [cited 2022 Aug 22];5(4):e227067. Available from: https://doi.org/10.1001/jamanetworkopen.2022.7067

4. Greenhalgh J. U.S. Life Expectancy Fell By 1.5 Years In 2020, The Biggest Drop Since WWII. NPR [Internet]. 2021 Jul 21 [cited 2022 Aug 22]; Available from: https://www.npr.org/sections/coronavirus-live-updates/2021/07/21/1018590263/u-s-life-expectancy-fell-1-5-years-2020-biggest-drop-since-ww-ii-covid

5. Arias E, Tejada-Vera B, Kochanek KD, Ahmad FB. Provisional Life Expectancy Estimates for 2021. National Center for Health Statistics (U.S.), editor. 2022 Aug 31;(23). Available from: https://stacks.cdc.gov/view/cdc/118999

6. NVSS - Provisional Death Counts for COVID-19 - Executive Summary [Internet]. 2022 [cited 2022 Oct 29]. Available from: https://www.cdc.gov/nchs/covid19/mortality-overview.htm

7. Noymer A, Garenne M. The 1918 Influenza Epidemic’s Effects on Sex Differentials in Mortality in the United States. Popul Dev Rev [Internet]. 2000 [cited 2022 Aug 28];26(3):565–81. Available from: https://www.ncbi.nlm.nih.gov/pmc/articles/PMC2740912/

8. Goldstein JR, Lee RD. Demographic perspectives on the mortality of COVID-19 and other epidemics. Proceedings of the National Academy of Sciences [Internet]. 2020 Sep 8 [cited 2022 Aug 24];117(36):22035–41. Available from: https://www.pnas.org/doi/abs/10.1073/pnas.2006392117

9. Vaupel JW. Life Expectancy at Current Rates vs. Current Conditions: A Reflexion Stimulated by Bongaarts and Feeney’s “How Long Do We Live?” Demographic Research [Internet]. 2002 Aug 15 [cited 2022 Oct 17];7(8):365–78. Available from: https://www.demographic-research.org/volumes/vol7/8/

10. Vaupel JW, Manton KG, Stallard E. The impact of heterogeneity in individual frailty on the dynamics of mortality. Demography [Internet]. 1979 Aug [cited 2016 Apr 19];16(3):439–54. Available from: http://link.springer.com/article/10.2307/2061224

